# Capturing infant and child growth dynamics with P-splines mixed effects models

**DOI:** 10.1101/2025.10.22.25338570

**Authors:** María Alejandra Hernandez, Zheyuan Li, Tim J Cole, Yi Ying Ong, Kate Tilling, Ahmed Elhakeem

## Abstract

Investigating early life growth dynamics is crucial for a more comprehensive understanding of the developmental origins of obesity. Spline methods based on basis splines (B-splines) provide excellent flexibility for modelling complex nonlinear growth patterns, but they are prone to overfitting. To ensure good fit and avoid overfitting, B-splines can be extended by adding a penalty term to control their flexibility, resulting in what are commonly known as penalized B-splines (P-splines). Despite their strengths, P-splines are not yet widely used in epidemiology, partly due to a lack of practical guidance. This paper provides an illustrative guide to using P-spline linear mixed effects models to examine early life growth trajectories and estimate key growth features in longitudinal studies. After detailing P-spline theory and model fitting, we apply the method to repeated measurements of height, weight, and body mass index (BMI) up to age 10 years in a Southeast Asian birth cohort. We estimated infant growth velocity, and magnitude and timing of infant peak BMI and childhood rebound BMI, and explored sex differences, intercorrelations, and associations with prenatal factors. In our cohort, infant peak growth velocity was higher in boys than girls, ages of peak and rebound BMI had a negligible correlation, and greater birth length was associated with lower infant height velocity and higher weight velocity. We discuss practical considerations, alternative modelling approaches and provide recommendations for research. P-splines simplify the knot selection process, making them a valuable approach for growth modelling. R library, code and datasets are provided to accelerate uptake.

## INTRODUCTION

Infant and childhood growth trajectories vary between individuals, are primarily shaped by genetic factors and prenatal environments, and can influence subsequent development and disease risk^1-3^. Growth is commonly assessed using anthropometric measures such as height, weight, and body mass index (BMI: a weight to height ratio). Height reflects skeletal growth; weight encompasses both lean mass and fat mass; and BMI provides an indirect measure of fat mass^4^. Trajectory modelling, i.e., the analysis of repeated measures over time, is crucial for understanding growth patterns and examining their determinants and effects on health^5,6^. Moreover, it enables the description of key growth features that are vital for understanding developmental dynamics^7,8^. For example, the timing of the childhood BMI rebound can identify children at increased risk of later obesity^9,10^.

Appropriately modelling complex nonlinear growth requires the use of advanced methods. Splines are flexible, state-of-the-art tools for capturing such patterns. They come in various forms, primarily distinguished by their polynomial degree (e.g., linear versus cubic spline)^11-13^. Conventional regression splines are widely used for longitudinal analyse but they can be prone to overfitting because they rely onto manual knot selection^11,14^. P-splines address this limitation by applying a penalty to balance between model fit and smoothness^14,15^. However, despite their conceptual advantages and established utility for growth chart development^16^, the use of P-splines for longitudinal growth curve analysis in epidemiology remains limited. This likely stems from a perceived complexity and lack of accessible software and guides, which stresses a need for clear guidance to encourage adoption in epidemiological studies.

This paper addresses this need by providing a practical guide to using P-spline linear mixed effects models to characterise infant and child growth curves to estimate key growth features in longitudinal studies. We begin by providing an overview of the linear mixed effects model. Next, we introduce P-splines as a flexible tool for nonlinear curve fitting, explaining how P-splines extend classic B-splines through penalisation to ensure appropriate smoothing while avoiding excessive flexibility. We then describe the P-spline linear mixed effects model for curve fitting and derivative estimation for predicting growth features. Following this, the modelling and estimation process is illustrated on data from the Growing Up in Singapore Towards healthy Outcomes (GUSTO) Study. Lastly, we discuss practical considerations, alternative modelling strategies, and recommendations for methodological research.

## NONLINEAR GROWTH CURVE MODELLING WITH P-SPLINES

### Linear mixed effects (LME) model

Mixed effects models, also known as random effects or multilevel models, are widely used for analysing repeated measures data. Mixed effects models include fixed effects to estimate a population-average (mean) growth curve and random effects to capture individual-specific deviations from this overall average curve. One common approach is the linear mixed effects (LME) model, which models the outcome as a linear combination of the fixed and random effects^17,18^. An LME model for a single continuous repeated outcome (e.g., weight) as a linear function of a single continuous predictor (e.g., age or time) that includes random intercepts and random slopes, is written as follows:

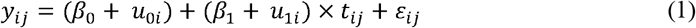

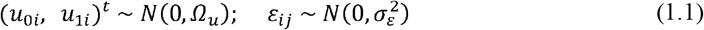

where *y*_*ij*_ denotes a single outcome measured in the i-th individual (*i* = 1,2, …, *N*) at age *t*_*ij*_ (j = 1,2, …, *J*_*j*_), with the responses *y*_*ij*_ assumed to be independent between individuals. The parameters *β*_*o*_ and *β*_1_ are fixed effects that represent the average intercept and average slope across all individuals in the population, respectively. The terms *u*_*oi*_ and *u*_1*i*_ are random effects that quantify how much the intercept and slope for the i-th individual deviate from the population-average intercept and slope, respectively. The random effects are assumed to be normally distributed with mean zero and covariance matrix *Ω*_*u*_. This assumption ensures that the fixed effects represent the population-level average. The residual error *ε*_*ij*_ is assumed to be independently and identically normally distributed with mean zero and variance 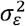. Random effects and residual errors are assumed to be mutually independent.

### P-splines LME model

The LME model in Eq. (1) assumes that the outcome changes linearly with age. This model can be extended to capture nonlinear change by replacing the linear age term with nonlinear function of age (e.g., a global quadratic polynomial function for age), with the LME model remaining linear in its coefficients, meaning the relationship between the predictors and the outcome is modelled through a linear combination of model parameters. Among the most flexible tools for modelling nonlinear patterns are splines, which can be integrated into the mixed effects modelling framework to replace the linear age term with a curve defined by piecewise polynomials^11,13^.

A widely used and computationally efficient choice is to represent the smooth curve using B-spline basis functions, which provide numerical stability due to their local support and their flexible properties^11,19^. A spline curve expressed in B-spline form is a linear combination of several B-spline basis functions of degree *q*. Each basis function is a piecewise polynomial defined over *q* + 1 adjacent polynomial segments that are smoothly connected at *q* specified points called knots. The knots for the full set of basis functions are distributed across the age range, and the combined curve retains continuity of derivatives up to order *q* − 1 at the knots. A common choice is the cubic B-spline which uses degree-3 polynomials (*q* = 3) and gives a smooth cubic spline curve with continuous 1^st^ (velocity) and 2^nd^ (acceleration) derivatives.

The flexibility of a cubic spline is determined by the number (and, to lesser extent, location) of the knots (**Figure 1**). The number and location of knots must be set manually by the user and this choice can have a significant impact on the B-spline fit. Using too many knots risks overfitting, where the spline becomes excessively ‘wiggly’ by modelling the noise alongside the underlying pattern. This reduces model generalisability and increases uncertainty (wider confidence intervals)^11,14^. Selecting the optimal knot structure is a well-known challenge with no single-best solution. A common strategy is to compare models with different knots using information criteria such as the Bayesian Information Criterion (BIC). However, this is only valid for Maximum Likelihood (ML) estimation and might not always identify the most appropriate model. This reliance on manual knot selection undermines reproducibility. An alternative family of functions known as penalized splines mitigate these issues and simplify the process of knot selection by imposing a roughness penalty on the spline^20^.

**Figure 1.**
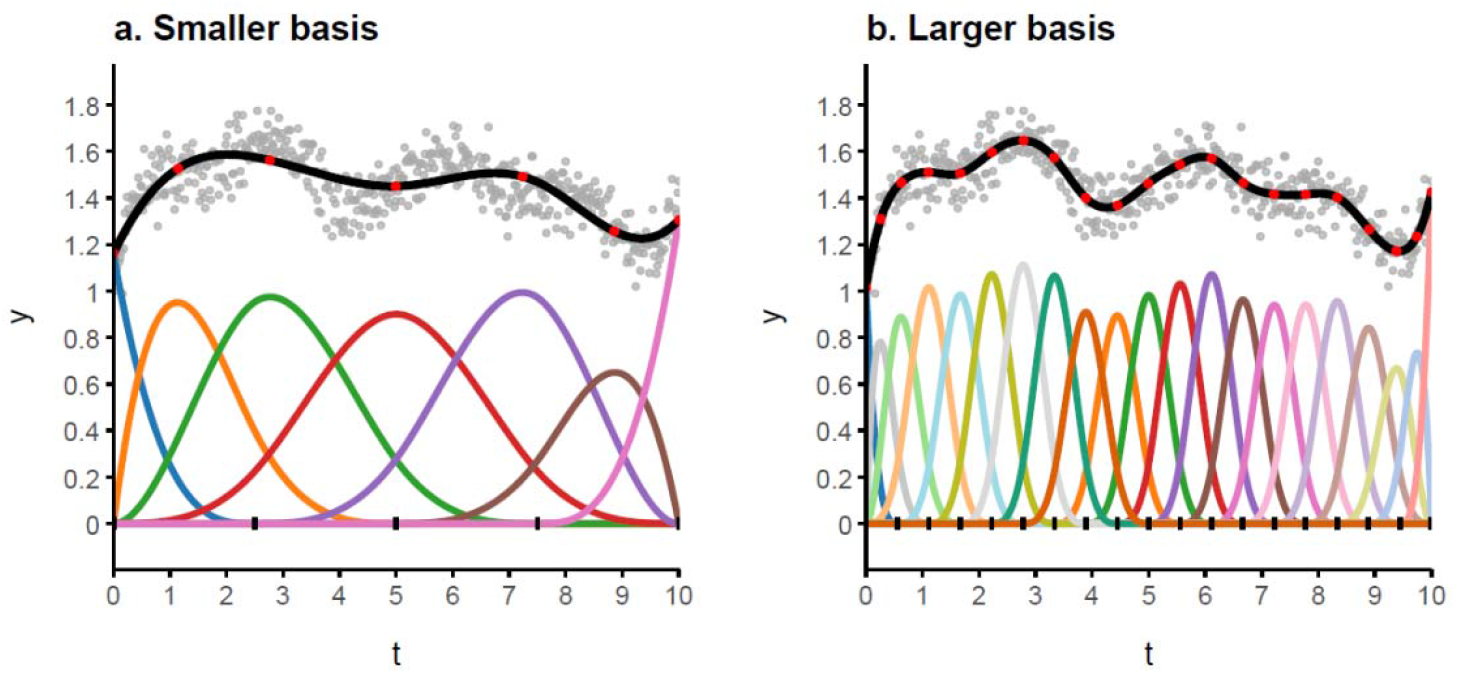
Cubic B-spline construction and the impact of basis size on spline flexibility. Figure shows 2 cubic B-splines with evenly spaced knots fitted to the same simulated noisy data (grey points). The left panel (a) uses 7 basis functions (3 internal knots) and the right panel (b) uses 21 (17 knots). Fitted splines are shown in black. Coloured curves represent the individual cubic B-spline basis functions, each scaled by its fitted coefficient from the linear model, which together form the spline. The small black ticks across the horizontal axis indicate the locations of the knots, which are the points where the spline’s polynomial segments are smoothly joined. Red points on the spline mark its value at the x-coordinate where each corresponding basis function reaches its peak, marking the region of strongest influence of that basis function on the spline. Comparing panels illustrates that increasing basis size provides greater local control and pattern capture but also increases the risk of overfitting by capturing random noise instead of underlying trends.

Penalized B-splines, known as P-splines, introduced by Eilers and Marx^14^, are a popular type of penalized splines. They consist of two components: first, a set of uniformly spaced knots; and second, a discrete difference penalty of order *p*, which enforces smoothness by smoothing across adjacent B-spline basis coefficients^14,15^. This penalty helps to prevent overfitting and makes the number of knots less critical. Instead of having to carefully select the number of knots, the user specifies a large approximate basis dimension that will provide sufficient flexibility to capture the underlying trend. The penalty then reduces the influence of the final basis size by effectively shrinking the coefficients of the basis functions, allowing the model to find a balance between fitting the data and remaining smooth.

To fit a flexible model within the mixed effects framework that captures a nonlinear relationship between the outcomes and the age covariate, we define the cubic P-spline LME model presented by Djeundje and Currie^21^ for the outcome of the i-th subject at the j-th measurement time *t*_*ij*_ as follows:

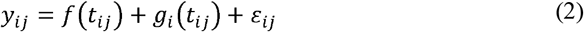

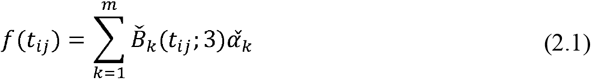

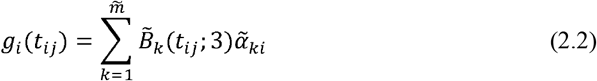

Where 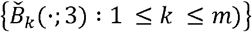 is a set of cubic B-spline basis functions representing the population function *f* (i.e., the fixed effects); 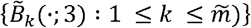 is a set of 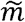 cubic B-spline basis functions representing the individual-specific deviations *g*_*i*_ (i.e., random effects); and 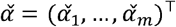 and 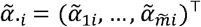 are two coefficient vectors for the fixed and random B-splines, respectively. To complete the full P-spline specification of the model in Eq. (2), we define two *p*-th order difference penalties for *f* and *g*_*i*_ as:

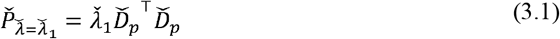

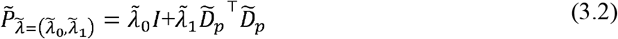

As we are working with discrete penalties, *Ď*_*p*_ and 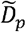 are constructed as *p*-th order difference matrices to discourage large jumps between adjacent basis coefficients. A common choice for enforcing smoothness is the second-order difference (*p* = 2). Allowing different values of *p* for each smooth term in the model e.g., using *p* = 2 for *f* and *p* = 1 for *g*_*i*_, can help reduce collinearity between the population-level and individual-level smoothers^22,23^. 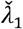 and 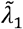 are two smoothing parameters that determine the strength of the penalty on the fixed 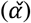 and random 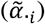 basis coefficients, respectively. 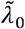 has an additional ridge penalty *I* that addresses identifiability issues^21^.

Selecting the optimal value for the smoothing parameter (*λ*) is critical because it controls the weight of the penalty. When *λ* = 0, the fit reverts to the B-spline curve in Eq. (2). Increasing the value of *λ* produces progressively smoother curves by shrinking coefficient differences closer to zero. As *λ* approaches infinity, the fit nears a straight-line estimate. One useful approach for estimating *λ* in our setting is to transform the P-spline into a (linear) mixed effects model^22,24,25^, where the (unpenalized) bases are expressed as fixed effects components and *λ* as a ratio of variance components (variance of residual errors to variance of the random effects). When specified in this way, *λ* can be automatically estimated from the data as part of the model fitting process with standard methods like restricted maximum likelihood (REML).

The `mgcv` library in R_20_ is a leading tool used for fitting various penalized splines, including penalized splines mixed models. However, `mgcv` (along with its related function `gamm()` and the associated `gamm4` library) relies on dense model matrices, limiting its ability to efficiently process large datasets (e.g., >1,000 individuals). In contrast, the `lme4` library^26^, widely used for fitting LME models, uses sparse matrices, enabling scalability, but has limited support for incorporating penalized splines. To address this gap, we developed the `psme` library^27^, which combines the strengths of `mgcv` and `lme4` to efficiently fit the P-spline LME model. Briefly, `psme` constructs the B-spline basis and penalty matrices in `mgcv` and reparametrizes these matrices into fixed and random effect components, which are then assembled into sparse matrices and fitted as an LME model in `lme4`.

### Derivative curve estimation

Derivative estimation provides critical insights into growth dynamics by quantifying growth velocity (the first derivative of a growth curve) and acceleration (second derivative)^7,24^. After fitting a P-spline LME model to repeated anthropometric data, derivatives for the population and individual-specific smoothing functions can be obtained by differentiating the B-spline basis functions and multiplying them by their estimated coefficients. The derivative curves can be used for characterising growth patterns, including peak growth velocity. In addition, derivative turning points (where 1^st^ or 2^nd^ derivative changes sign) can be used to identify landmarks on the growth curve such as the infant peak and child rebound BMI (**Figure 2**).

**Figure 2.**
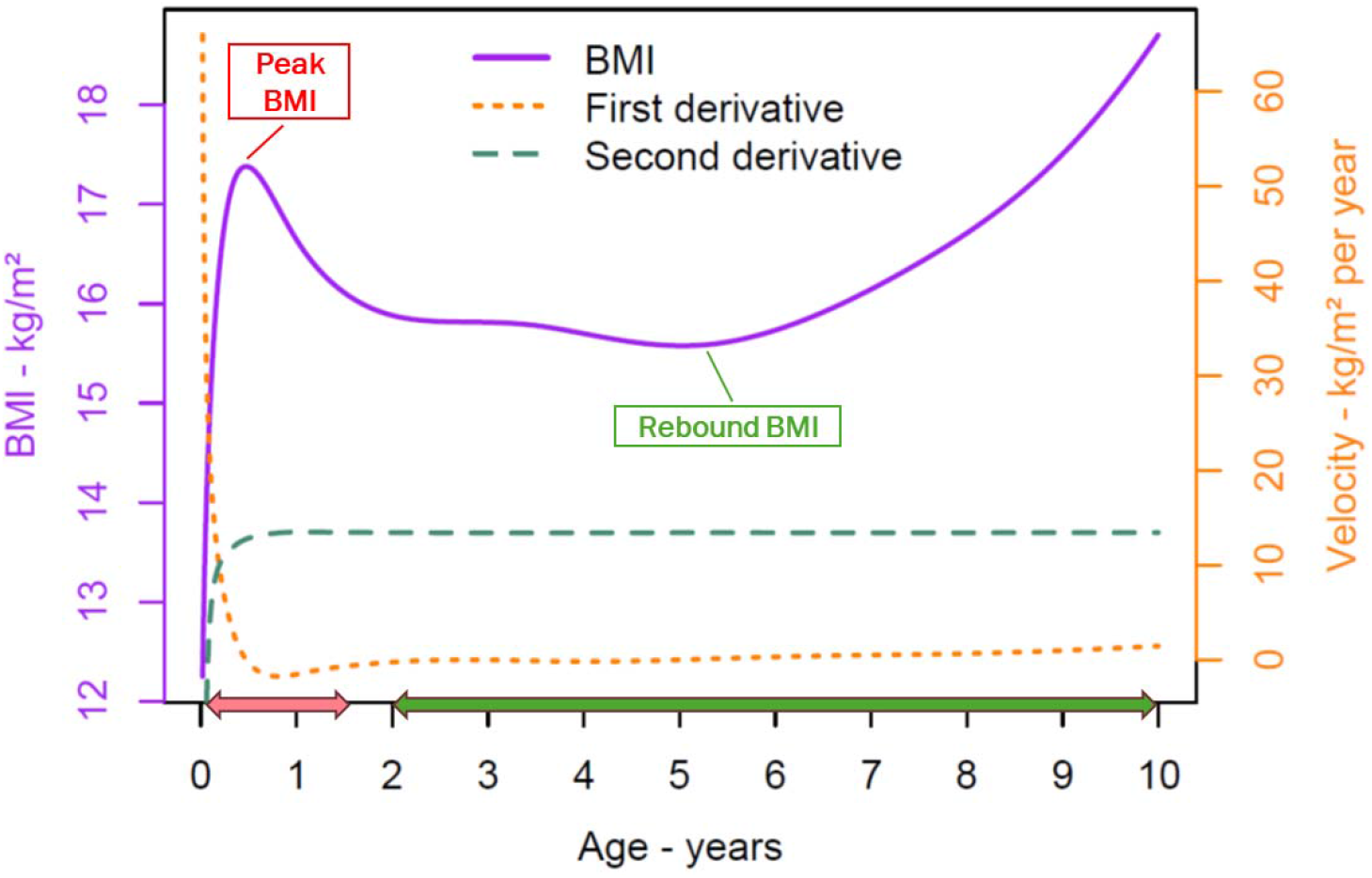
Schematic illustrating the infant peak BMI and childhood rebound BMI. Figure shows the infant peak BMI and childhood rebound BMI, along with the derivatives which will be used to estimate these BMI features from the fitted P-spline LME models. The second derivatives is scaled for illustration to allow it to fit in the figure. Infant peak BMI (*kg/m*^*2*^) is defined as the highest BMI during the first rise of the BMI curve up to age 18 months. Rebound BMI in *kg/m*^*2*^ marks the 2^nd^ rise in BMI after the infant peak and is defined as the lowest BMI after age two years. Peak and rebound BMI can be calculated by identifying the points where the BMI acceleration (second derivative) changes sign. The timings are based on the corresponding ages at peak and rebound BMI, expressed in months and years respectively.

## THE GUSTO STUDY

### Study design and participants

Pregnant women attending their first trimester antenatal ultrasound scan at Singapore’s two major public maternity units between 2009 and 2010 were invited to participate in GUSTO^28^. Women were eligible if they were aged 18 years or older, Singaporean citizens or permanent residents, with self-reported homogeneous ethnic ancestry (Chinese, Indian, Malay), intended to deliver at either of the recruitment hospitals and reside in Singapore for the next five years.

Women >14 weeks gestation, receiving chemotherapy, psychotropic medications, or with an existing type I diabetes mellitus diagnosis at recruitment, and those who did not agree to donate birth tissues (cord, placenta, and cord blood) were excluded. Out of 2,034 eligible women invited to participate, 1,450 enrolled in the study at 7-11 weeks of gestation. From this group, 1,344 women with a singleton, naturally conceived pregnancy were included in the main GUSTO cohort, with 1,098 mother-infant pairs remaining in the main GUSTO cohort at the time of delivery.

### Growth measurements

Up to 20 repeated measurements of length/height (cm) and weight (kg) were collected by trained staff during research clinics at approximate ages 3, 6, 9, 12, 15, 18, 24, 36, and 48 months, and 4.5, 5, 5.5, 6, 6.5, 7, 8, 9, and 10 years. Exact age (days) at each measurement was recorded. Crown-to-sole (recumbent) length was measured up to age 24 months (Seca 210 Mobile Measuring Mat) and standing height was measured from age 18 months (Seca 213 Stadiometer). Since both length and height were measured at 18 and 24 months, we prioritised length measurements at these ages, and if length was missing, height was used (n=70 at 18 months; n=74 at 24 months). Weight was measured by calibrated Seca 334 or Seca 803 digital scales. BMI was calculated as weight (kg) divided by height (m^2^).

Of the initial 1,098 infants, 1,039 had at least one height and weight measurement recorded between one week and ten years of age (N=15,554 observations). Erroneous values such as duplicates, biologically implausible measurements, carried-forward entries, and extreme outliers were identified and removed using the `growthcleanr` R library^29^, which led to the exclusion of 346 observations. Children with only a single measurement were then excluded, giving a final growth modelling study sample of 1,014 children (48% girls) with at least two repeated measurements of height, weight, and BMI from one week to ten years (N=15,183 observations). Each child contributed a median of 17 (interquartile range: 4) measurements. The height, weight, and BMI data used for growth modelling are presented in **Figure 3**.

**Figure 3.**
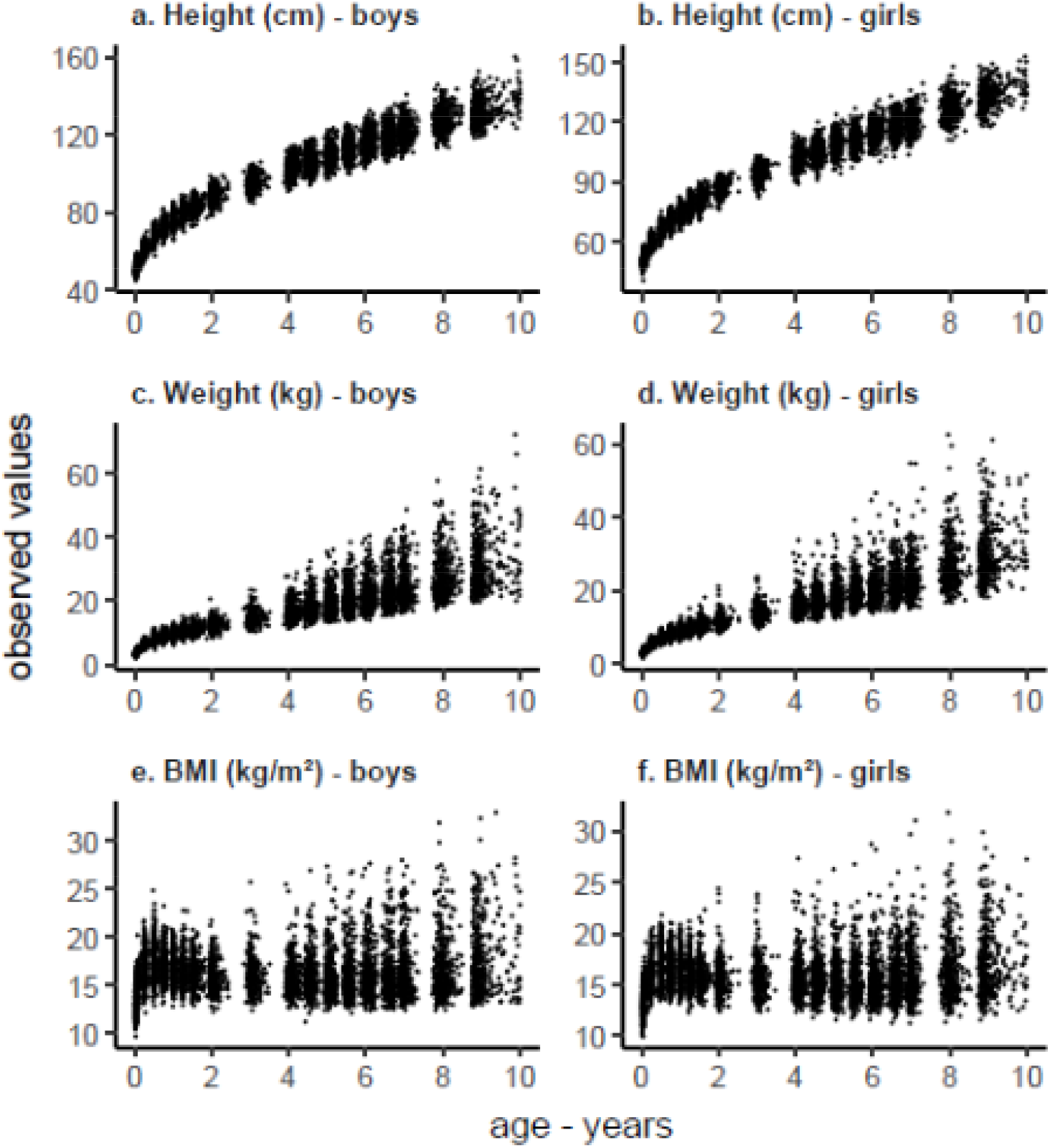
Observed height, weight, and BMI measurements in the GUSTO study. Figure shows the 15,183 longitudinal observations of height, weight, and BMI from 1,014 GUSTO boys and girls that were used for P-spline LME analysis. The data span from age one week to 10 years and each individual had a minimum of two repeated measurements.

### Growth curve modelling and estimation of growth features

The P-spline LME model for age described in Eq. (2) was used to construct population (fixed effects) and individual-specific (as random effects) weight, height, and BMI trajectory curves from age one week to ten years. P-splines were set up as cubic B-splines with evenly spaced

knots and bases of size 10 for both the functions *f* and *g*_*i*_. The basis size of 10 is larger than the number of knots used in other growth modelling studies^30-33^ and should provide sufficient flexibility to capture the nonlinear growth patterns while remaining computationally efficient.

We applied a 2^nd^ order difference penalty on adjacent bases for the fixed effects spline *f* and a 1^st^ order difference penalty on the random spline *g*_*i*_. Models were fitted separately in boys and girls due to expected sex differences in growth trajectories. A square root transformation was applied to age to improve spline fit by better accommodating the rapid growth at younger ages^15,34^. Models (and *λ*) were estimated via REML and fitted using the `psme` library in R^27^.

We generated the predicted mean and individual-specific growth curves for height, weight, and BMI by evaluating the B-splines on an age grid of 550 points (corresponding to weekly intervals from one week to ten years) and combining them with the model coefficients. This was done using the `EvalSmooth()` and `xTraj()` functions from the `psme` library. We also generated height and weight velocity curves by calculating the first derivative of the predicted growth curves.

BMI and age at the infant peak and childhood rebound BMI were estimated by identifying trajectory turning points using predicted BMIs up to age 18 months and from age 2 years, respectively. We first identified the approximate location of turning points by calculating where the smoothed second derivative changes sign: a change from positive to negative indicated a peak, and change from negative to positive indicated a trough (rebound). The location of these turning points was subsequently refined by iteratively fitting a quadratic model to an expanding window of neighbouring data points until a well-defined parabolic curve was established. BMI and age at peak and rebound BMI were calculated from the vertex of this final parabola. These calculations were done using the `getPeakTrough()` function from the `sitar` library^35^. A similar approach was used to estimate infant peak height and weight velocity from smoothed first derivatives (up to age 12 months).

### Follow-up analysis of estimated growth Features

We summarised the distribution of growth features using boxplots and assessed interrelations with Pearson correlation coefficients. Subsequently, we used linear regression to examine the associations of six prenatal and birth factors (maternal age, weight, and height, and offspring gestational age at birth, birth weight, and birth length) with growth features. The prenatal and birth factors were obtained from maternal reports, clinical measurements, or birth records and were standardised to mean=0, standard deviation (SD)=1 prior to analysis. Models were fitted in boys and girls combined with adjustment made for sex.

### Additional growth metrics

Lastly, to illustrate other measures that can be used to study growth dynamics, we calculated and described the infant height and weight growth velocity at specific ages (1, 6, 12, and 24 months) and compared height and weight trajectories up to age 5 years against World Health Organization (WHO) standards. For the latter, height-for-age and weight-for-age differences were calculated as the difference between predicted values and WHO median at ages 1, 6, 12, 18, 24, 30, 36, 42, 48, 54, and 60 months^36^. Predicted mean height-for-age and weight-for-age differences were estimated using random intercepts LME models with a natural spline for age plus an age-by-sex interaction to explore sex differences in growth patterns.

### Ethics

Study protocols followed the principles of the Declaration of Helsinki and were approved by respective ethics committees for two hospitals: National Healthcare Group Domain Specific Review Board (NUH) and SingHealth Centralized Institutional Review Board (KKH). All participants provided informed consent or assent to participate and contribute their data to publications.

### Computer code and synthetic dataset

All analyses were performed in R version 4.5.0 (R Project for Statistical Computing) and RStudio 2025.09.0+387 (Posit Software). All R code and a synthetic copy of the GUSTO dataset are available on GitHub at https://github.com/LongitudinalModelling/psplines. The synthetic growth data are provided to allow interested readers to practice the methods and were generated from P-spline LME model predictions via the `synthpop` library in R^37,38^.

## Results

Predicted height, weight, and BMI growth curves are shown in **Figure 4**, and were consistent with the observed data and literature on early life growth patterns^1^. **Figure 5** plots predicted growth curves against observed weight, height, and BMI for six randomly selected children with different numbers of repeated measurements and shows that model predictions closely mirror the observed data for all outcomes with no qualitative difference by the number of repeated measurements. The predicted height and weight velocity curves are presented in **Figure 6** and show the expected rapidly decelerating growth in early life^1^.

**Figure 4.**
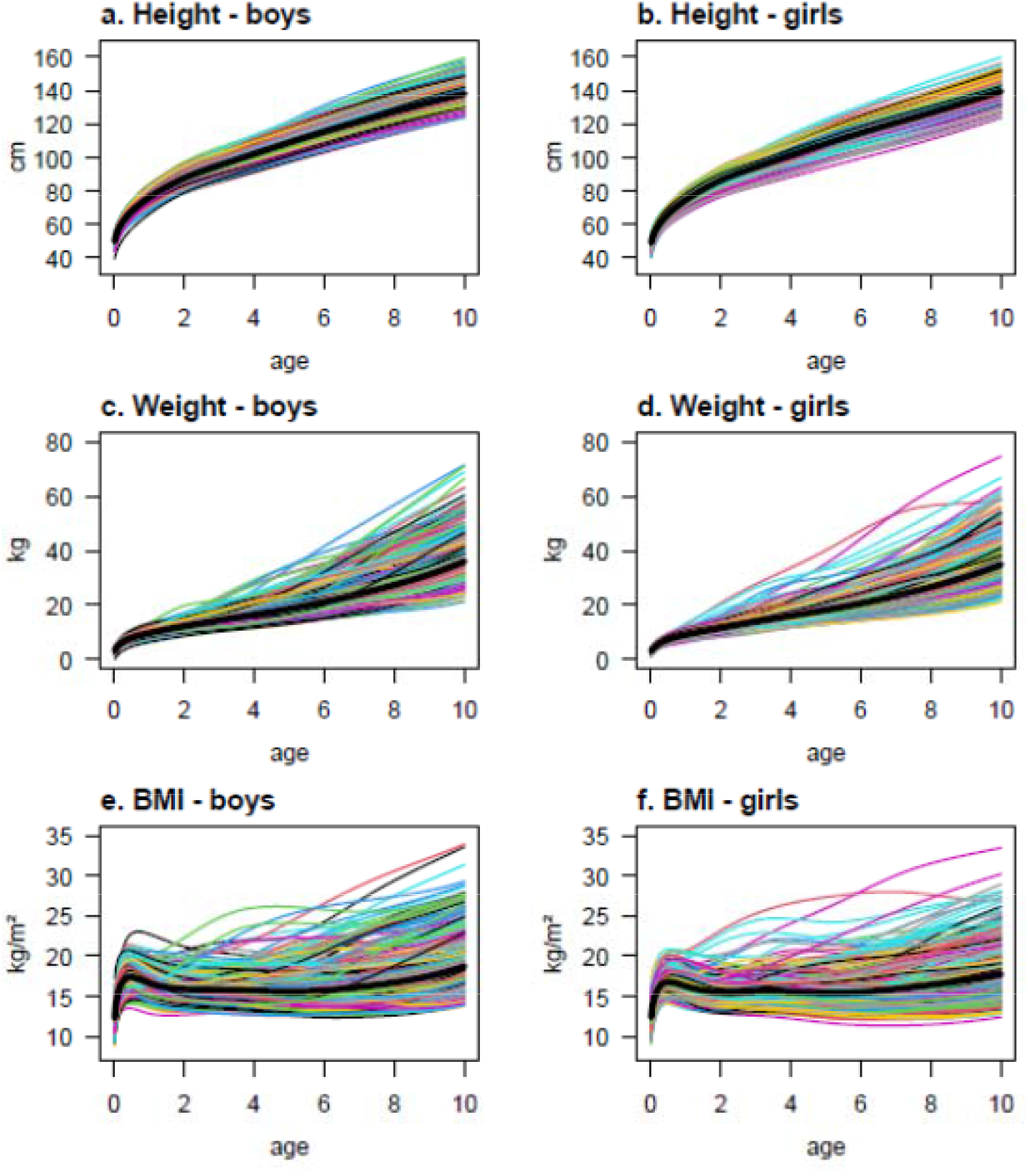
Predicted height, weight, and BMI growth curves. Figure shows predicted height, weight, and BMI growth curves from age one week to ten years from P-spline LME models in GUSTO boys and girls. Coloured lines are the individual-specific growth curves; black lines are population-average growth curves.

**Figure 5.**
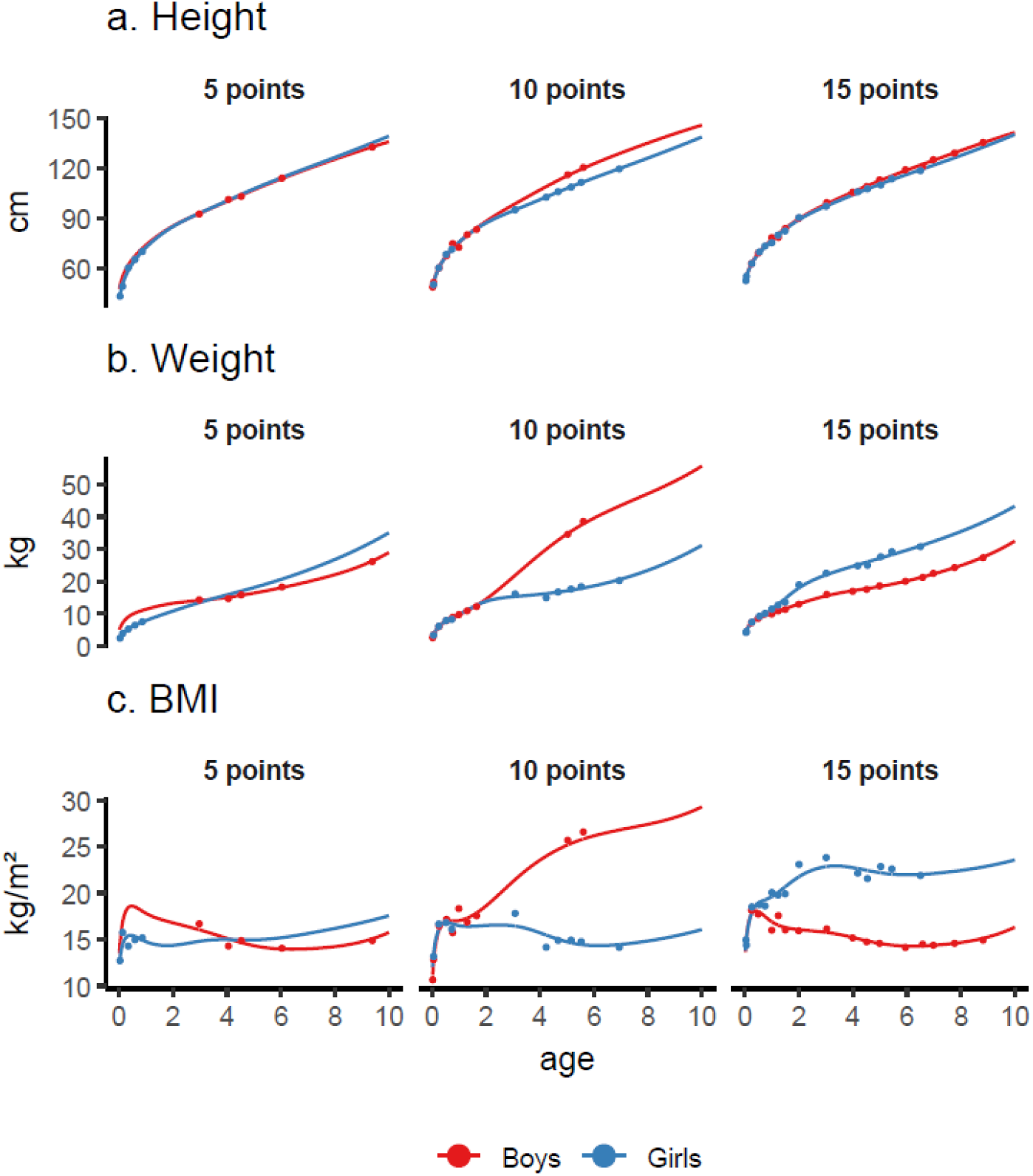
Predicted versus observed weight, height, and BMI for a randomly selected boy and girl with respectively 5, 10, and 15 repeated growth measurements. Figure shows predicted growth curves (from P-spline LME models) versus the observed values (circles) for three boys and three girls that were randomly selected according to the number of repeated measurements (5, 10, or 15).

**Figure 6.**
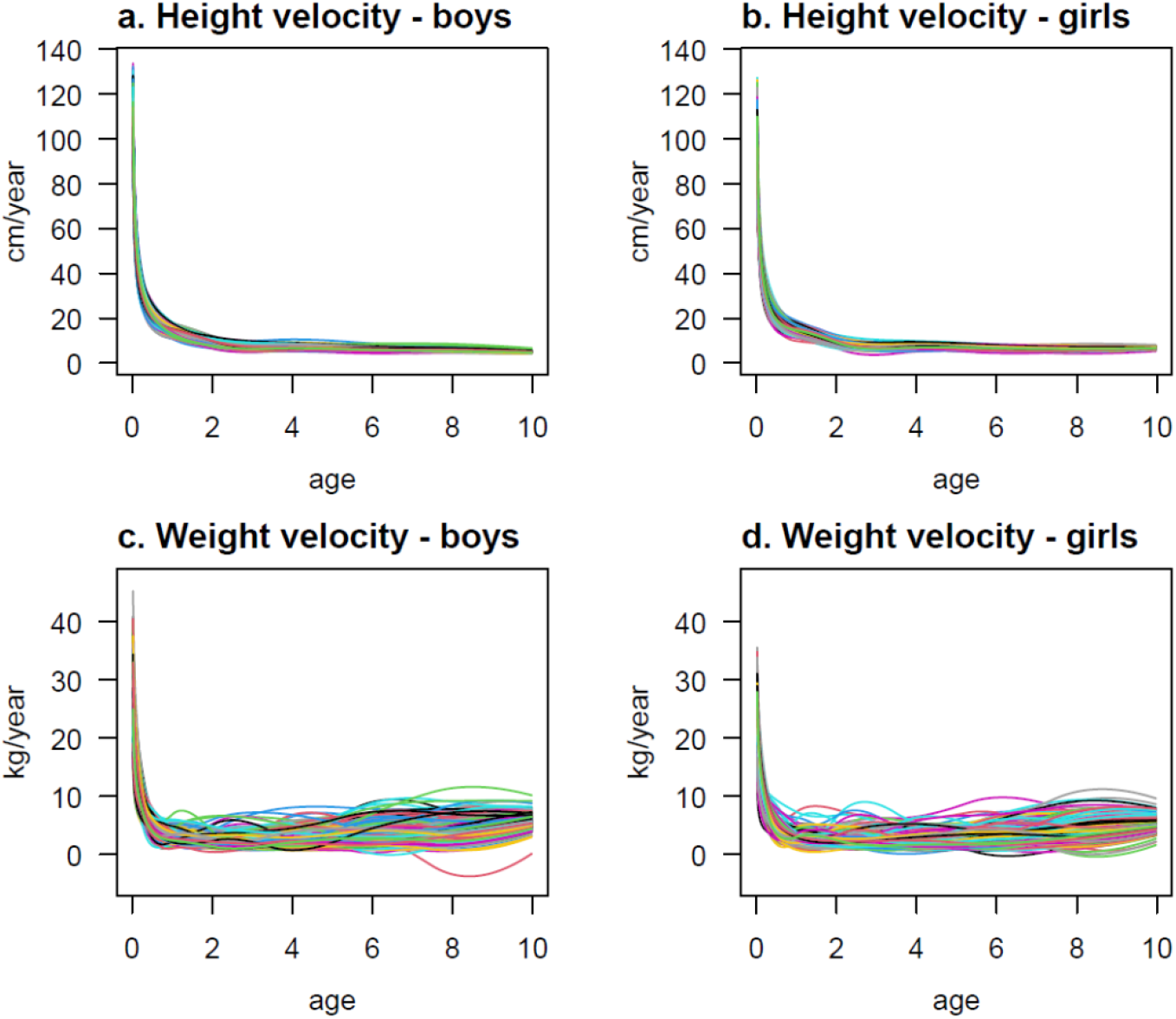
Predicted weight and height velocity curves. Figure shows the individual-specific height and weight growth velocity curves from 1 week to 10 years in GUSTO boys and girls (i.e., the first derivative of the predicted growth curve).

Of the 1,014 children included in the P-spline LME modelling, 109 (10.7%) had no peak or rebound BMI (no peak BMI: n=22, no rebound BMI: n=82, neither: n=5), and 169 (16.7%) had no peak height or weight velocity (no height velocity: n=161, no weight velocity: n=6, neither: n=2). **Figure 7** shows boxplots of the estimated growth features by sex. On average, boys (vs. girls) had a higher infant peak height velocity (median:.4.3 vs. 3.9 *cm/month*) and a higher peak weight velocity (1102 vs. 882 *grams/month*), but estimates were more similar for peak BMI (17.4 vs. 16.8 *kg/m*^*2*^), rebound BMI (14.9 vs. 14.7 *kg/m*^*2*^), age at peak BMI (5.6 vs. 6.1 *months*), and age at rebound BMI (5.5 *years*). There was also considerable variability in growth features between children (**Figure 7**). For example, SD around the mean infant peak height velocity in boys (mean: 4.4 *cm/month*) was 0.3, and the SD around the mean age at rebound BMI in girls (mean: 5.4 *years*) was 1.6. BMI and age at rebound BMI estimates were consistent with the literature^9,10^. Our BMI and age at peak BMI estimates were very similar to previous estimates (obtained using different methods) in GUSTO^30,39^.

**Figure 7.**
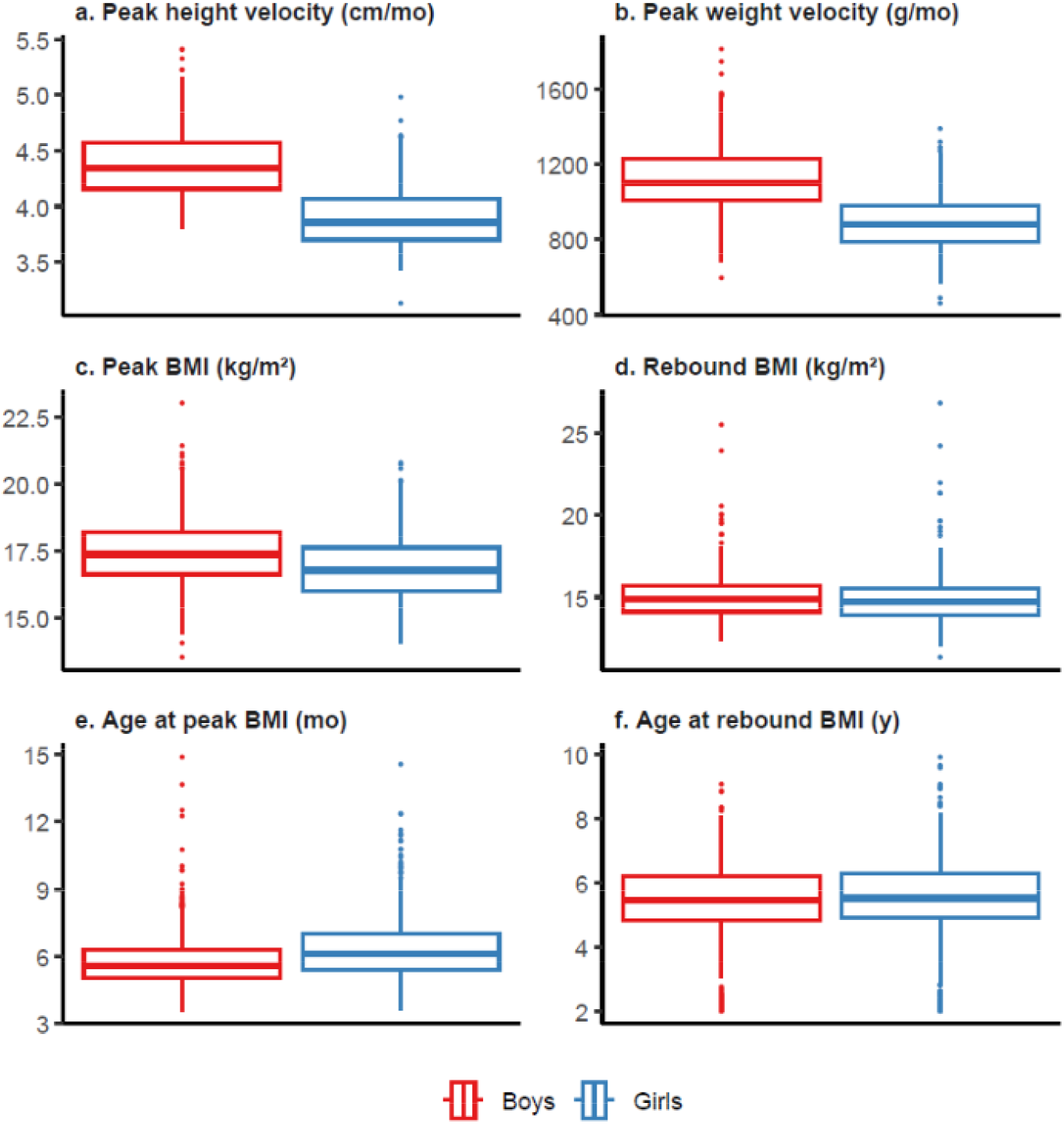
Distribution of growth features. Figure shows box plots summarising distribution of infant peak height velocity (cm/month) infant peak weight velocity (grams/month), the magnitude (kg/m^2^) and timing (months) of infant peak BMI, and the magnitude (kg/m^2^) and timing (years) of childhood rebound BMI in boys and girls in the GUSTO cohort. Each box represents the middle 50% of distribution (i.e., interquartile range). The line inside the box is the median. Whiskers (lines extending from the top and bottom of the box) extend to the minimum and maximum values within 1.5 times the interquartile range. Data points that fall outside the whiskers are plotted as individual points. Means (SD) in boys and girls were 4.4 (0.3) and 3.9 (0.3) [peak height velocity: cm/month], 1121 (181) and 890 (150) [peak weight velocity: grams/month], 17.4 (1.3) and 16.8 (1.2) [peak BMI: kg/m^2^], 15.1 (1.4) and 14.9 (1.5) [rebound BMI: kg/m^2^], 5.8 (1.3) and 6.4 (1.5) [age at peak BMI: months], 5.4 (1.4) and 5.4 (1.6) [age at rebound BMI: years].

Pairwise correlations between the growth features are shown in **Figure 8** and were virtually identical in boys and girls. These included a negligible correlation between the ages of peak and rebound BMI (*r* ≤ 0.02), low negative correlation between the BMI and age at rebound BMI (*r* ≤ -0.27), low positive correlation between infant peak height and weight velocity (*r* ≤ 0.31), and moderate positive correlations between peak and rebound BMI (*r* ≤ 0.57), and between infant peak weight velocity and peak BMI (*r* ≤ 0.70).

**Figure 8.**
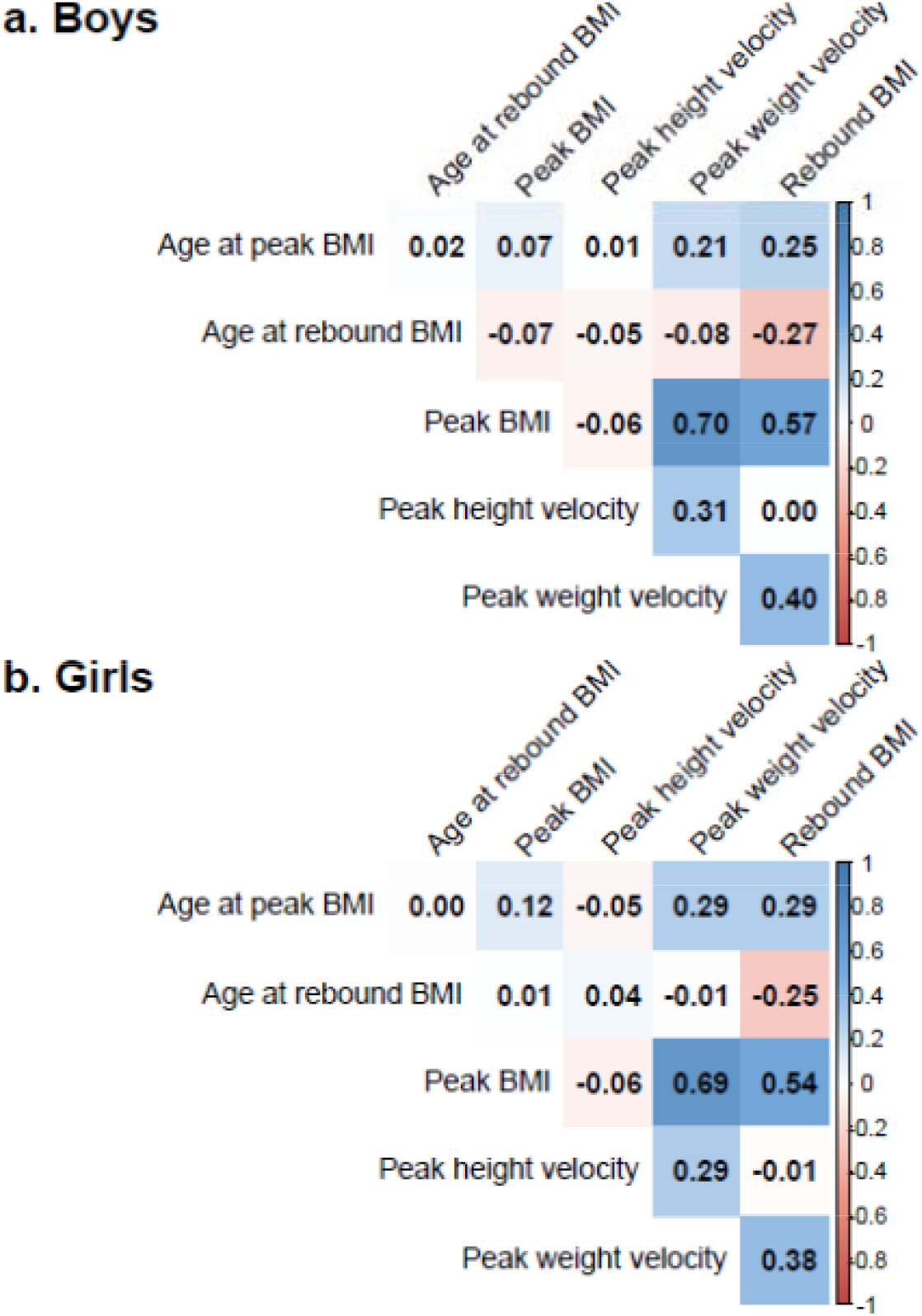
Growth feature correlations. Figure shows correlation matrices for infant peak height/weight velocity and magnitude and timing of the infancy peak BMI and childhood rebound BMI, separately in boys and girls from the GUSTO study. The numbers represent Pearson correlation coefficients for pairs of growth features.

Figure 9 presents the associations of maternal prenatal and birth factors with infant and child growth features. Among maternal factors, higher maternal weight was associated with higher rebound BMI and a younger age at rebound BMI. Higher maternal height was associated with higher peak height and weight velocity, and higher peak and rebound BMI. Maternal age was not associated with growth outcomes. For example, mean difference in rebound BMI per SD unit higher maternal weight was 0.26 *kg/m*^*2*^ (95%CI: 0.17 to 0.35). Regarding birth factors, higher gestational age was associated with lower infant peak height velocity, younger age at peak BMI, and higher rebound BMI. Higher birth weight and birth length were associated with lower infant peak height velocity, higher infant peak weight velocity and higher peak and rebound BMI. Higher birth weight was also associated with younger age at peak BMI. For example, mean difference in infant peak height velocity per SD unit higher gestational age was -0.69 *cm/month* (-0.93 to -0.44).

**Figure 9.**
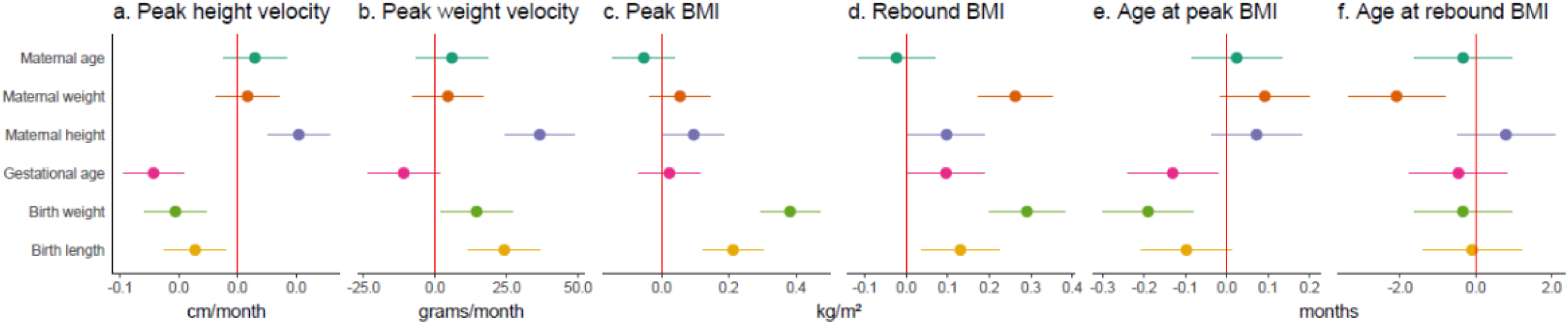
Association of maternal prenatal and birth factors with infant and child growth features. Figure shows mean differences (points) and 95% confidence intervals (horizontal bars) in infant peak height and weight growth velocity and the magnitude and timing of the infant peak BMI and childhood rebound BMI per a standard deviation unit higher maternal age, maternal weight, maternal height, gestational age, birth weight, and birth length in the GUSTO cohort. Models included both boys and girls and were adjusted for sex. Mean differences in age at rebound BMI are shown in months to aid presentation. The analysis sample comprised 823 children with complete data on all variables.

Figure 10 presents boxplots and correlations of age-specific growth velocity (at 1, 6, 12, and 24 months). The figure shows that growth velocity is highest at 1 month and that correlations between velocities is strongest for adjacent time points and progressively weakens as the time interval increases, with almost no correlation between the 1-month and 24-month velocities. Mean height-for-age and weight-for-age differences from age 1 month to 5 are presented in **Figure 11** and suggest that the study participants’ height was below the WHO median from age 30 months onwards.

**Figure 10.**
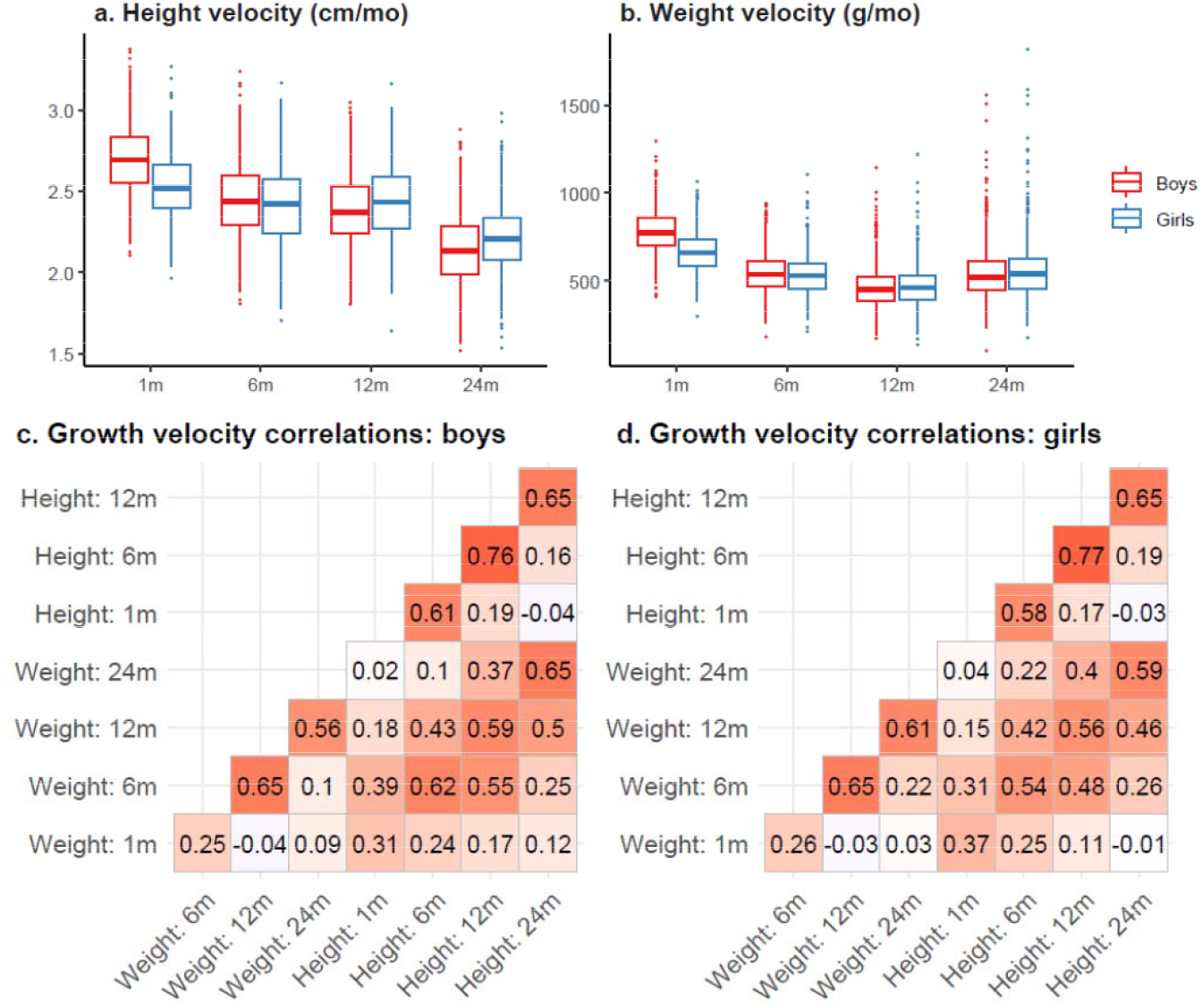
Distribution and correlations of infant height and wright growth velocity at ages 1, 6, 12, and 24 months. Figure shows box plots (a, b) and correlations matrices (c, d) for infant height and weight velocity at ages 1, 6, 12, and 24 months in boys and girls in GUSTO.

## DISCUSSION

### Summary

We introduced the P-spline LME model and demonstrated its application to early life growth trajectories in a South Asian birth cohort. P-splines address the challenge of knot selection by combining B-splines with evenly spaced knots and applying a penalty term to reduce the risk of overfitting. This constistutes an improvement over conventional regression splines which require manual specification of both the number and location of knots. The fitted trajectories closely matched the observed data and existing literature, supporting validity of the P-spline LME approach. We also illustrated how curve derivatives can be estimated, highlighting the model’s utility in deriving critical growth metrics that offer novel insights into developmental processes. For example, the negligible correlations between growth velocity at ages 1 and 24 months suggests that infant growth during the first month may be more strongly determined by intrauterine environments, and that genetics become more influential later in infancy to guide children towards their predetermined growth trajectory, which supports the concept of homeorhesis (or canalization)^1^.

**Figure 11.**
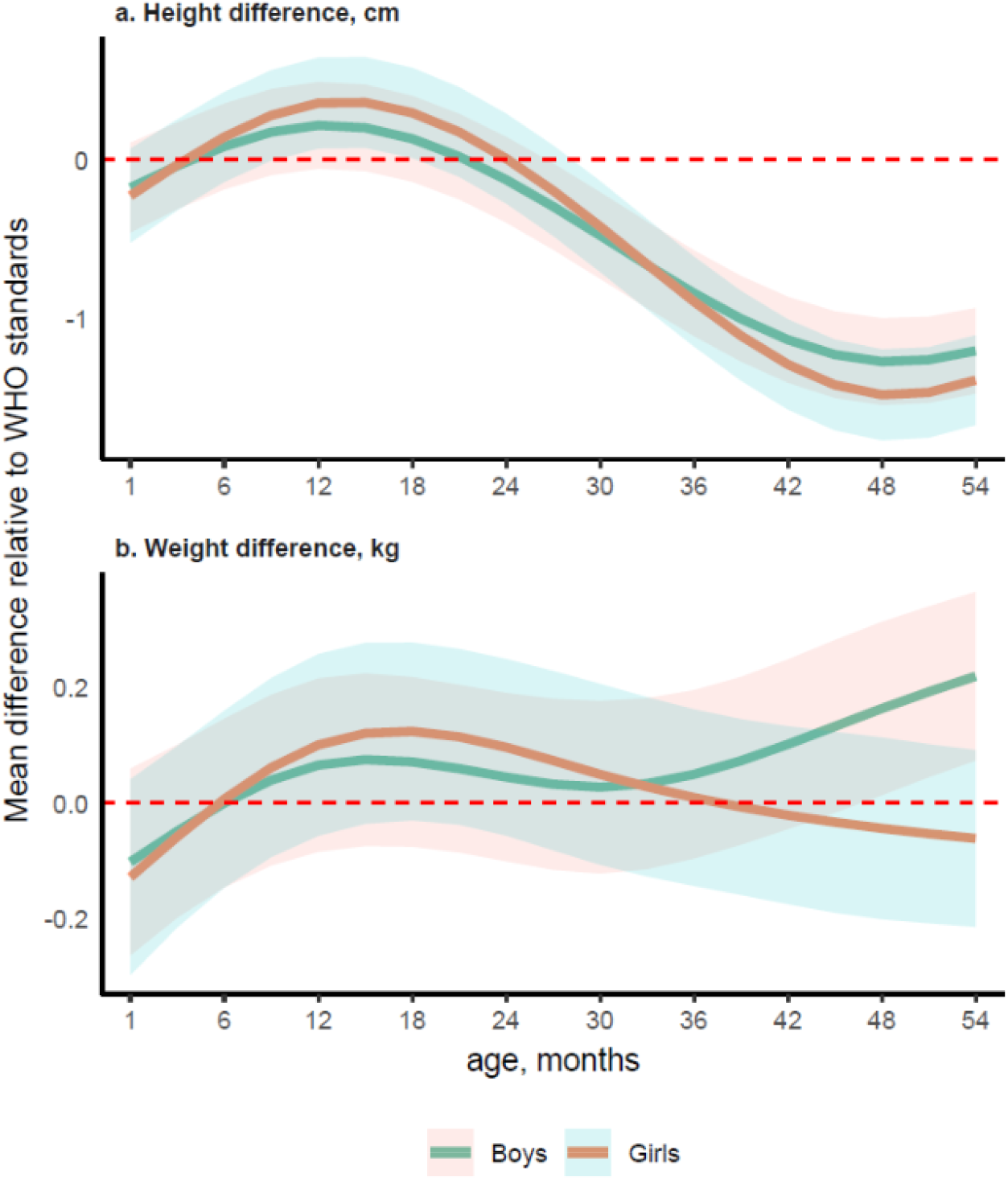
Mean height and weight difference relative to WHO standards from age 1 month to 5 years. Figure shows the mean difference (lines) in height-for-age and weight-for-age up to age 5 years relative to WHO medians at each age, by sex. Height-for-age and weight-for-age differences were calculated by subtracting predicted height and weight (from P-spline LME models) from the WHO median at each age. Analysis was done using linear mixed effects models that included a natural spline (3 knots) for age plus its interaction with sex, and random intercepts. The shaded area represents 95% confidence intervals.

### Practical considerations

Our `psme` library reformulates P-splines as mixed models with sparse fixed and random effects to enable efficient estimation. It took only 34 seconds to fit the six P-spline LME models, highlighting the `psme` library’s efficiency and scalability for larger datasets. The mixed model representation also supported REML (or ML) estimation of the smoothing parameters. We chose REML instead of ML estimation because ML estimates of variance components tend to be biased downwards, which can lead to oversmoothing^40,41^. REML is also preferred over generalized cross-validation because it tends to produce smoother fits^42^.

We used 1^st^ order differences penalty for the random spline so that it is not collinear with the population spline (2^nd^ order). Choice of penalty order can impact the flexibility and shape of the fitted curve and so it is worth exploring different options^15^. The penalty in `psme` follows Djeundje and Currie^21^ but alternatives have been proposed^42^. One such method adds a second difference penalty to both the population and individual-specific curves to improve derivative estimates^43^. Applying this double penalty to GUSTO, and optimising smoothing parameters using the Separation of Overlapping Precision Matrices method^24,44^, showed that the growth features derived from the double penalty approach were strongly correlated with those from our `psme` penalty method (**Figure 12**).

**Figure 12.**
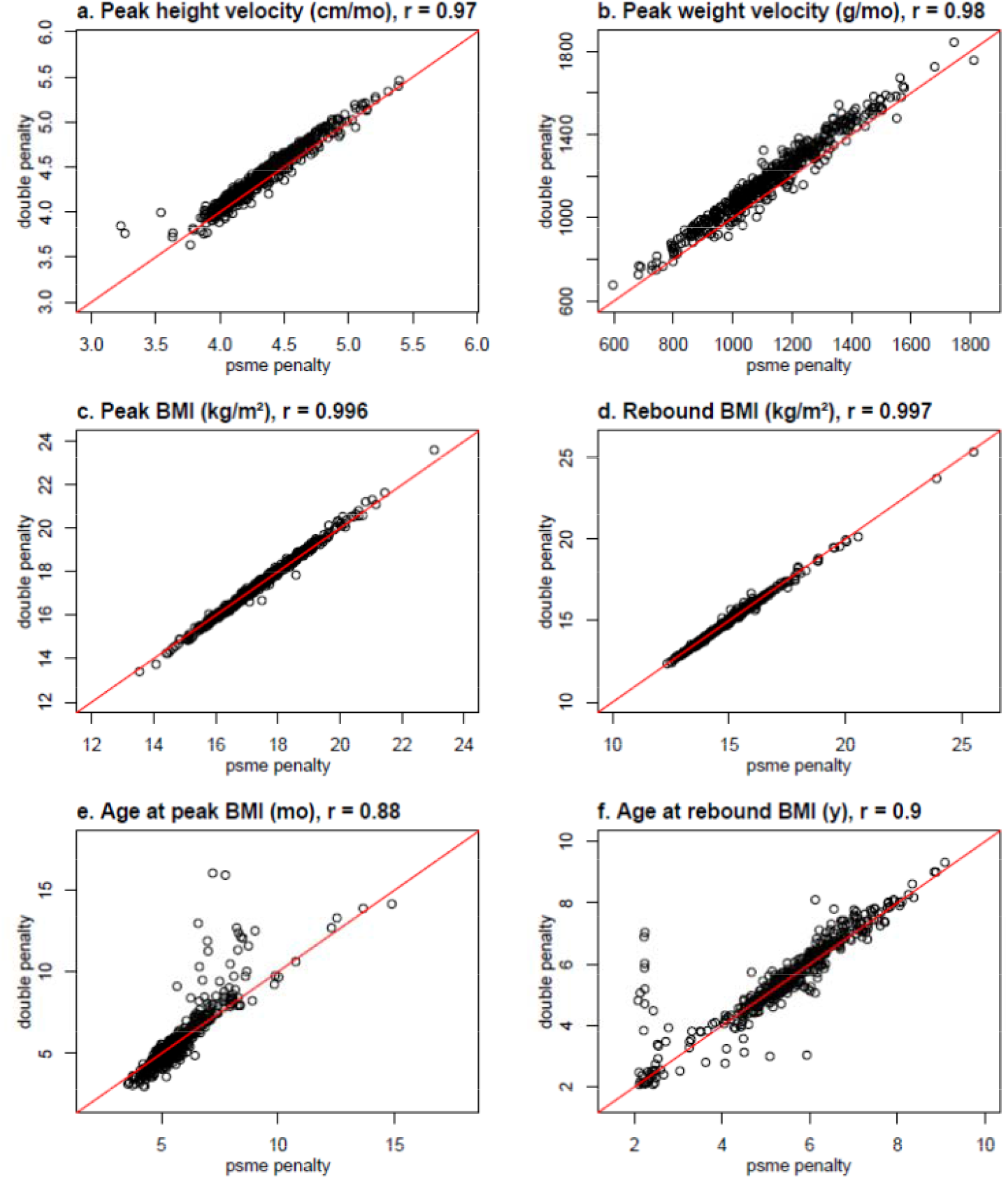
Comparison of growth features estimated from our P-spline models (psme) with estimates from P-spline models using the double penalty specification. Figure shows scatterplots and Pearson correlation coefficients (r) between growth features estimated from P-spline LME models that use the penalty method in the psme library (i.e., the models used in this paper) and P-spline LME models using the double penalty method. The red lines are lines of equality. Data are shown for GUSTO boys.

Following the classic P-spline approach of Eilers and Marx^14^, we selected a B-spline basis rather than alternatives such as truncated power functions^45^, which are considered inferior due to poor numerical conditioning and an inflexible penalty structure^46^. Using 10 bases provided good flexibility and computational efficiency and should be sufficient for most applications in longitudinal trajectories in epidemiological studies. It is generally impossible, at least theoretically, to have too many B-splines because the penalty rather than the number of knots controls the smoothness. However, increasing basis sizes comes with computational cost. Further, we used the same basis size for both population and random splines but studies with fewer repeated measurements may need to use a smaller basis in the random spline.

Transforming the age covariate can improve model fit by linearising relationships^15,34^, and in our example, a square root transformation of age produced more suitable curves than either the original age scale or a log transformation. Transforming the repeated outcome can help address skewness and heteroscedasticity to ensure residuals better meet model assumptions, but such transformations alter the scale and interpretation of the estimated parameters. It is worth noting that 10.7% of GUSTO participants had no identifiable peak or rebound in BMI, which may reduce the precision of estimates in subsequent analyses and their exclusion can distort associations with later outcomes^47^.

### Alternative modelling approaches

Numerous alternative methods exist for modelling growth trajectories. Functional principal component analysis, for example, estimates the mean trajectory using a functional basis and captures individual-specific deviations through principal components^48-50^, often smoothing mean and covariance functions with P-splines^51^ or local polynomials^52^. Another widely used method, SITAR (Super Imposition by Translation And Rotation), models a mean curve with a natural spline and summarises individual-specific curves using three random effects^53,54^.

LME models with fractional polynomials can also be used^55,56^ as can global polynomials^57^ though the latter lack the local support found in splines. Linear spline LME models are also commonly used, and while they do not produce smooth curves, they provide interpretable slope estimates for specified age windows^13,58-60^. Structural equation modelling approaches such as latent change score models can also be useful^61^. At the group level, clustering methods like latent trajectory models can identify subgroups with different trajectories^13,62^. More recently, approaches that integrate machine and deep learning within mixed effects and functional data frameworks are emerging as promising for prediction tasks^63-65^. Finally, P-splines can also be implemented within a Bayesian framework^66^.

### Recommendations for methodological research

Research is needed to systematically evaluate P-spline LME models against other approaches to growth curve analysis and in cohorts with differing numbers of repeated measures, sample sizes, age ranges, and patterns of missing data. Comparative studies are also needed to assess the impact of different P-spline model specifications, including alternative age and outcome transformations and penalty structures. Studies that demonstrate the utility of P-splines for modelling pooled individual-level data from multiple cohorts, and for modelling outcomes beyond physical growth such as mental health score trajectories, are also warranted.

### Final remarks

P-spline LME models are a valuable method for examining nonlinear growth curves in longitudinal population studies. The `psme` library, R scripts to replicate analyses, and a synthetic GUSTO dataset are provided to support uptake of this method. We hope this resource enables epidemiologists to apply the method in their own longitudinal studies.

## Data Availability

The data used are described in https://gustodatavault.sg. Interested researchers may request for data by contacting the corresponding author. Research proposals and data release will be assessed by the GUSTO Executive Committee.

## ABBREVIATIONS

BMI: body mass index
B-spline: basis spline
GUSTO: Growing Up in Singapore Towards healthy Outcomes
LME: linear mixed effects
ML: Maximum Likelihood
P-spline: Penalized B-spline
psme: Penalized Splines Mixed Effects Models
REML: restricted maximum likelihood estimation
SITAR: Super Imposition by Translation And Rotation
SD: Standard deviation
WHO: World Health Organization

## Availability of data and materials

R code and synthetic data can be found https://github.com/LongitudinalModelling/psplines.

## Acknowledgments

We thank the GUSTO study group and all clinical and home-visit staff involved. The voluntary participation of all participants is greatly appreciated. The GUSTO study group includes. Allan Sheppard,Amutha Chinnadurai, Anne Ferguson-Smith, Anne Eng Neo Goh, Arijit Biswas, Audrey Chia, Birit Leutscher-Broekman, Borys Shuter, Shirong Cai, Cheryl Ngo, Chai Kiat Chng, Shang Chee Chong, Christiani Jeyakumar Henry, Mei Chien Chua, Cornelia Yin Ing Chee, Yam Thiam Daniel Goh, Dennis Bier, Chun Ming Ding, Doris Fok, Eric Andrew Finkelstein, Fabian Kok Peng Yap, George Seow Heong Yeo, Wee Meng Han, Helen Chen, Hugo P S Van Bever, Hazel Inskip, Iliana Magiati, Inez Bik Yun Wong, Jeevesh Kapur, Jenny L Richmond, Jerry Kok Yen Chan, Joshua J Gooley, Krishnamoorthy Niduvaje, Bee Wah Lee, Yung Seng Lee, Leher Singh, Sok Bee Lim, Lourdes Mary Daniel, Seong Feei Loh, Yen-Ling Low, Pei-Chi Lynette Shek, Marielle Fortier, Mark Hanson, Mary Foong-Fong Chong, Michael Meaney, Susan Morton, Wei Wei Pang, Pratibha Agarwal, Anqi Qiu, Boon Long Quah, Rob M van Dam, David Stringer, Salome Antonette Rebello, Wing Chee So, Chin-Ying Hsu, Lin Lin Su, Jenny Tang, Kok Hian Tan, Soek Hui Tan, Oon Hoe Teoh, Victor Samuel Rajadurai, PC Wong and Sudhakar K Venkatesh The funders had no role in study design, data collection and analysis, interpretation of data, or writing of this report.

## Contributors

AE developed the idea and study design, with input from MAH and ZL. AE did the analysis with input from MAH and ZL. MAH developed the double penalty approach. ZL developed the `psme` library. MAH and AE wrote the first manuscript draft. ZL, TJC, YYO, and KT provided comments on the draft and approved the final version.

